# What is known about the determinants of developing antipsychotic-induced metabolic syndrome and interventions to address them for community dwelling adults: a scoping review protocol

**DOI:** 10.1101/2023.01.05.22283663

**Authors:** Emma Good, Debi Bhattacharya, Michelle Maden, Sion Scott

## Abstract

1.0

**Introduction:** Antipsychotics are the primary treatment for severe mental health conditions. Whilst antipsychotics are effective at improving psychiatric outcomes, approximately 80% of people will experience metabolic side effects (MSEs), characterised by weight gain, lipid disturbance and glucose dysregulation. Antipsychotic-induced MSEs are associated with a two-fold increased risk of developing coronary heart disease and a five-fold risk of developing type 2 diabetes.

Gender, ethnicity, age, and family history are reported non-modifiable determinants of developing antipsychotic-induced MSEs and indicate who is at highest risk. However, it is not clear which determinants are most significant to inform targeting interventions for high-risk individuals.

Antipsychotics induce increases in calorie intake and sedentary behaviours. Behavioural determinants are modifiable and provide potential intervention targets, however, the extent to which they have been studied and targeted is unclear.

The literature describes the testing of non-pharmacological interventions to target behaviours. However, few studies report clinically significant attenuation of MSEs, and the preferred healthcare setting to deliver an intervention to prevent antipsychotic-induced MSEs is yet to be established.

**Methods and analysis:** This review will adhere to the Joanna Briggs Institute guidance for scoping reviews and the PRISMA-ScR checklist (Appendix 1). Relevant electronic databases will be searched. Qualitative, quantitative and mixed method study designs, and evidence syntheses will be considered. One reviewer will independently screen titles and abstracts, with a 20% sample checked by two reviewers. Full text screening will be undertaken by one reviewer with a 10% sample checked by another. Data will be extracted and synthesised to address the research objectives.

**Ethics and dissemination:** Ethics approval is not required. Findings will be disseminated through professional networks, conference presentations and publication in a scientific journal.

**medRxiv registration details:** DOI:10.1101/2023.01.05.22283663

**STRENGTHS AND LIMITATIONS OF THIS STUDY:**

- This study will be the first to systematically identify the knowledge gaps the in body of literature relating to the modifiable and non-modifiable determinants of antipsychotic-induced metabolic side effects (MSEs) in community dwelling adults, the non-pharmacological interventions to target behaviours that have previously been implemented to prevent and/or treat antipsychotic-induced MSEs and their success or non-success, and the preferred context of delivery of such interventions from the point of view of the individuals affected by antipsychotic-induced MSEs. Insights from these can highlight areas to inform further research in this area.
- This study will link modifiable determinants of antipsychotic-induced MSEs to the theoretical domains framework (TDF), which can be linked to a taxonomy of behaviour change techniques to inform components for a future intervention.
- The review will take a rigorous approach, adhering to the Preferred Reporting Items for Systematic Reviews and Meta-Analyses-Scoping Review (PRISMA-ScR) guidelines.
- Only literature published in English will be included. This could potentially limit the diversity of literature captured in the review.

## 3.0 INTRODUCTION

### 3.1 Background

Antipsychotics are the mainstay of treatment for severe mental health conditions and are extremely effective at improving psychiatric outcomes(1). Though people with severe mental illness already have a higher risk of premature mortality than the general population(2,3), this is exacerbated with the use of antipsychotics(4,5). Metabolic changes can occur rapidly after the commencement of antipsychotics, and approximately 40%-80% of people prescribed antipsychotics will experience weight gain, glucose dysregulation, and dyslipidaemia, collectively described as metabolic side effects (MSEs) (1,6–9). This cluster of metabolic side effects occurring together is referred to as ‘metabolic syndrome’(10). Antipsychotic-induced MSEs are associated with the development of hypertension, a two-fold increased risk of developing cardiovascular diseases such as coronary heart disease, and a five-fold increased risk of developing type 2 diabetes(1-2,4,10-11). Metabolic changes increase the risk of premature mortality in this population by 33%-55%(13).

Research has reported various non-modifiable determinants indicating which people are at risk of developing antipsychotic-induced MSEs, including gender, ethnic background, age, and family history of obesity(4,5,7,14–18). It is not apparent how important each of these factors are in determining which groups are most at risk of developing MSEs, therefore, it is not known if and how the individuals at highest risk are targeted with interventions to prevent or treat antipsychotic-induced MSEs. This scoping review will explore what is known about which groups within the population of community dwelling adults are most at risk of developing antipsychotic-induced MSEs, and what is known about their non-modifiable determinants and modifiable determinants.

Non-pharmacological interventions to prevent and treat antipsychotic-induced MSEs demonstrate limited clinically efficacy in terms of decreases in weight, waist circumference, and body mass index (BMI), or long-term positive effects on diet and physical activity(19). Studies testing non-pharmacological interventions have been limited by short follow-up periods thus the sustainability of any improvements in clinical outcomes is uncertain. Process evaluations report patient barriers to engaging with intervention components, including difficulty in using self-monitoring tools, burden of regularly recording progress, barriers to accessing ongoing support, and resistance to undergo physical health checks and monitoring(2,19–22). As antipsychotics induce changes in behaviour, such as increased calorie intake and sedentary behaviour, modifiable determinants, such as dietary choices and physical activity levels, can attenuate and even prevent the physiological manifestations of MSEs(19,23–25). Non-pharmacological interventions require significant behaviour change from individuals to prevent the development of MSEs, however, there is limited evidence to suggest which behavioural determinants have previously been targeted and which interventions have been underpinned by behaviour change theory. This may explain why previous interventions have reported limited efficacy. Whilst the outcomes of interventions have been widely reported, it is not clear from the literature what the most successful context for delivery is when targeting behavioural determinants. Due to the traumatic nature of living with severe mental illness, during episodes of crisis focus is given to short-term goals, for example, managing psychosis(20). As such, the most appropriate context in terms of preferred healthcare setting for delivery of non-pharmacological interventions and time proximity to the initial prescribing decision to target behavioural determinants is yet to be established. Previous research suggests that interventions should be integrated with the early stages of pharmacological treatment to prevent or reduce the adverse metabolic effects of antipsychotics(26). This would certainly target the rapid metabolic changes after commencement of antipsychotics, and is clinically most likely to be effective to intervene at this stage, however, it is not known if this would be feasible during a traumatic acute psychiatric phase. Previous research also suggests that longer interventions should be developed, or the relationship between the antipsychotic treatment and non-pharmacological interventions should be understood further(26). Research also suggests that affected individuals would like regular contact with healthcare providers and support with lifestyle changes without rigid targets(22,27). This scoping review will describe what is known about the delivery of non-pharmacological interventions and will summarise the preferred contexts for implementation.

### 3.2 Study rationale

The literature regarding antipsychotic-induced MSEs, and the reported non-pharmacological interventions to prevent or treat this syndrome, highlight key unknown factors in relation to behavioural changes required by the affected population to prevent the development of MSEs. A preliminary search did not identify any scoping or systematic reviews investigating this. Scoping reviews are useful for answering broad research questions, to comprehensively review a complex or heterogeneous body of literature, and to identify main concepts and knowledge gaps in the evidence(28,29). A scoping review was chosen for this study to systematically assess and synthesise knowledge regarding what is known about the population most affected by antipsychotic-induced MSEs, and the modifiable and non-modifiable determinants that puts these individuals at high risk of developing MSEs. The review will scope and describe the non-pharmacological interventions that have previously been implemented to prevent or treat antipsychotic-induced MSEs from a patient behavioural perspective, and the context in which they were delivered. The scoping review will summarise the key messages from the evidence and identify any gaps in the current knowledge to understand the preferred context for the delivery of interventions for the target population. This will facilitate in informing future research regarding the implementation of behavioural interventions to prevent antipsychotic-induced MSEs.

### 3.3 Aim and objectives

The aim of this scoping review is to describe what is known about the determinants of developing antipsychotic-induced MSEs and interventions to address them for community dwelling adults, specifically to describe what is known and the knowledge gaps regarding the:

1. Non-modifiable determinants of the target population of community dwelling adults developing antipsychotic-induced metabolic side effects.
2. Modifiable determinants of the target population developing antipsychotic-induced metabolic side effects.
3. Non-pharmacological interventions targeting patient behaviours that aim to prevent and/or treat antipsychotic-induced metabolic side effects.
4. Preferred context for delivery of non-pharmacological interventions targeting patient behaviours to prevent and/or treat antipsychotic-induced metabolic side effects.

## 4.0 METHODS AND ANALYSIS

This review title has been registered with medRxiv (DOI:10.1101/2023.01.05.22283663). The review is being conducted by a team comprised of academics and clinicians in the field of behavioural science. The review is scheduled to start in November 2023 and conclude in March 2023.

### 4.1 Defining the research question

By using additional ‘timing’ and ‘setting’ elements with the PICO framework(30), the PICOTS framework(31) will be used to capture additional important contextual factors(32). PICOTS will structure the research question and will facilitate searching by focusing on addressing the review’s objectives, prompting the selection of key search terms, to clearly identify the problem, interventions implemented, and outcome(33):

- Population: Adults prescribed antipsychotics.
- Intervention/Exposure: Non-pharmacological intervention.
- Comparison: Unrestricted.
- Outcome: Metabolic syndrome and its physiological manifestations.
- Timing: Short- and long-term quantitative and qualitative outcomes following antipsychotic prescription.
- Setting: Community context.

### 4.2 Scoping review design

The scoping review will be guided by the aims and objectives of the study to understand the evidence base. Scoping reviews systematically map evidence using the Preferred Reporting Items for Systematic Review and Meta-analysis Extension for Scoping Reviews (PRISMA-ScR) checklist to help develop understanding of the relevant items, main concepts, and terminology, which will be utilised in this review(29). This scoping review will also be guided by the steps for scoping reviews outlined by Arksey and O’Mally(34), Levac et al.(35) and the Joanna Briggs Institute (JBI)(28) to (a) identify the research question, (b) identify relevant studies, (c) select studies, (d) extract and chart the data, (e) collate, summarise and report the results, and (f) conduct a consultation exercise with stakeholders.

The Cochrane Handbook for Systematic Reviews of Interventions(36) and Technical Supplement for Searching for and selecting studies(37) will also be used to guide this scoping review. The Handbook and Technical Supplement provide guidance on how to plan the search process, designing the search strategy, adapting search strategies across different databases, and searching for and selecting studies.

Due to the iterative nature of scoping reviews, any refinements to the review objectives and methods as a result of learning will be described in the results manuscript(28).

### 4.3 Search strategy

A preliminary search was undertaken in MEDLINE to identify key terms to develop a full list of search terms. A full list of 20 terms were agreed by the research team to be included in the search strategy (see Appendix 2). The search strategy was developed with guidance from an information specialist.

The search strategy consists of two parameters: population (people prescribed antipsychotics), and outcome (MSEs). Following a scoping exercise of search terms to define the search strategy, the MeSH (Medical Subject Heading)/Emtree terms ‘Metabolic Syndrome’ ‘Antipsychotic Agents’, ‘Psychotic Disorders’, ‘Schizophrenia’, and ‘Bipolar Disorder’ are used in the search. These search terms are adapted for the databases that do not permit MeSH/Emtree terms and database-specific Boolean operators will be applied.

As scoping reviews use an iterative process, the search strategy will be amended and refined, as necessary, to conduct an additional search using all identified keywords to find relevant sources across all databases. Any amendments and discrepancies will be detailed and justified in the manuscript.

### 4.4 Identifying relevant literature

#### 4.4.1 Eligibility Criteria

Eligibility criteria have been developed in consultation with the research team. Eligible studies will include those that are published in peer-reviewed journals between inception (1965) and the search date (November 2023); in the English language; and describe original research. Literature reviews, case reports, and protocols/ongoing studies will be excluded. Evidence syntheses will be excluded, however, their reference lists will be reviewed for eligible studies. Eligible studies will report on community-dwelling adults who have developed antipsychotic-induced MSEs, and will report one or more of the review’s objectives:

- Non-modifiable determinants of developing antipsychotic-induced MSEs.
- Modifiable determinants of developing antipsychotic-induced MSEs.
- Non-pharmacological interventions that aim to prevent and/or treat antipsychotic-induced MSEs by changing patient behaviours.
- Preferred context for delivery of non-pharmacological interventions targeting patient behaviour to prevent and/or treat antipsychotic-induced MSEs).

No assessments of items’ quality will be made, as the purpose of this scoping review is to synthesise and describe the coverage of the evidence.

#### 4.4.2 Inclusion and exclusion criteria

Inclusion and exclusion criteria for the search strategy have been constructed to identify potentially relevant studies as follows.

##### Inclusion criteria

- Adult population (≥18 years old).
- People in outpatient or community settings.
- People prescribed antipsychotics to treat psychosis/severe mental illness.
- MSEs described using any known definitions.
- Non-modifiable determinants – e.g., age,, ethnic background, sex, baseline BMI at antipsychotic initiation.
- Modifiable determinants – e.g., diet, physical activity, weight, blood pressure, blood glucose, blood cholesterol.
- Non-pharmacological interventions to change patient behaviours – e.g., lifestyle, dietary, physical activity.

##### Exclusion criteria

- Articles not written or translated in English.

Table 1 shows the inclusion and exclusion criteria for studies, pertaining to the population, concept and context (PCC) framework recommended by JBI for inclusion criteria for scoping reviews(38).

**Table 1:**
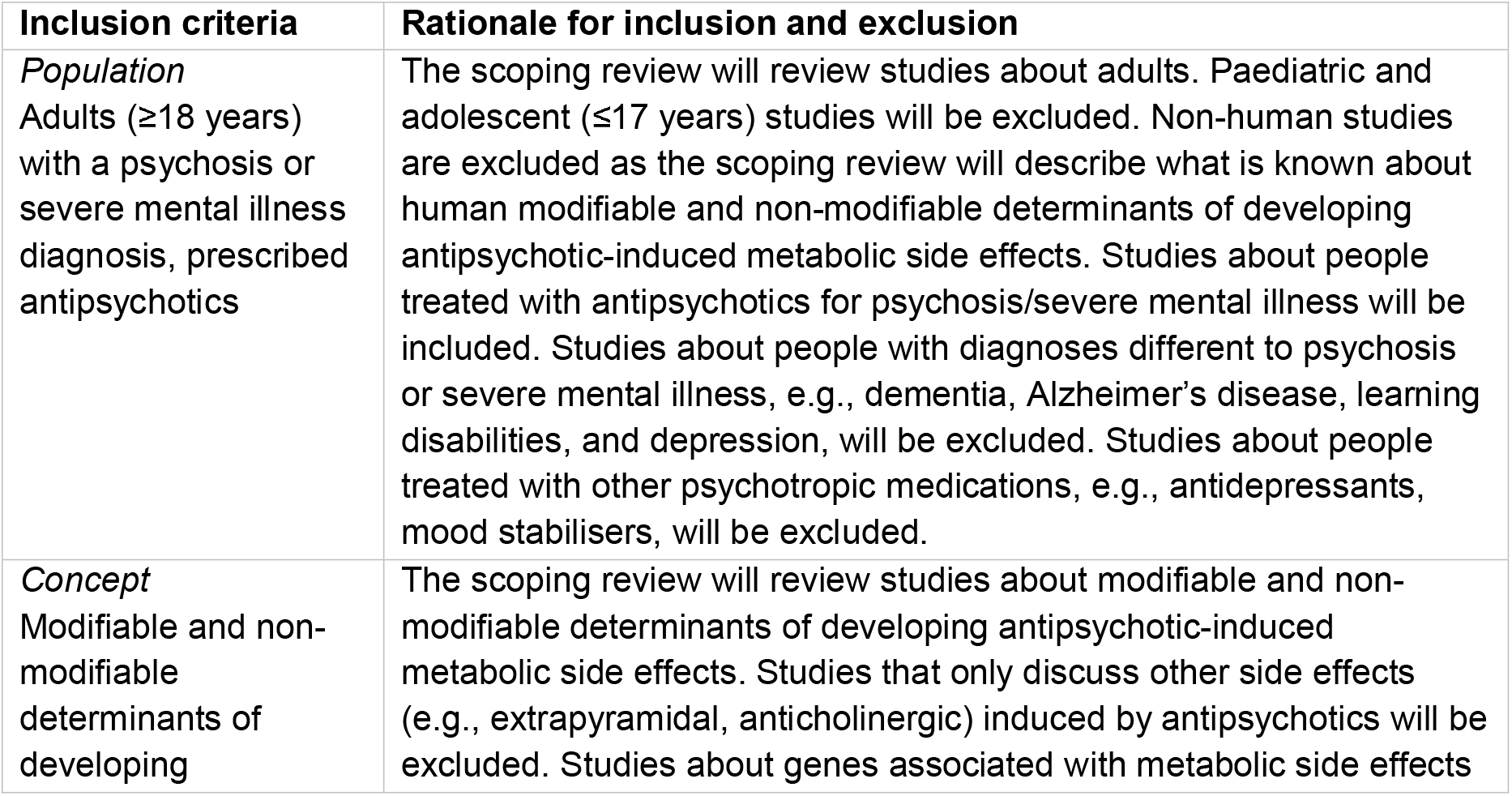
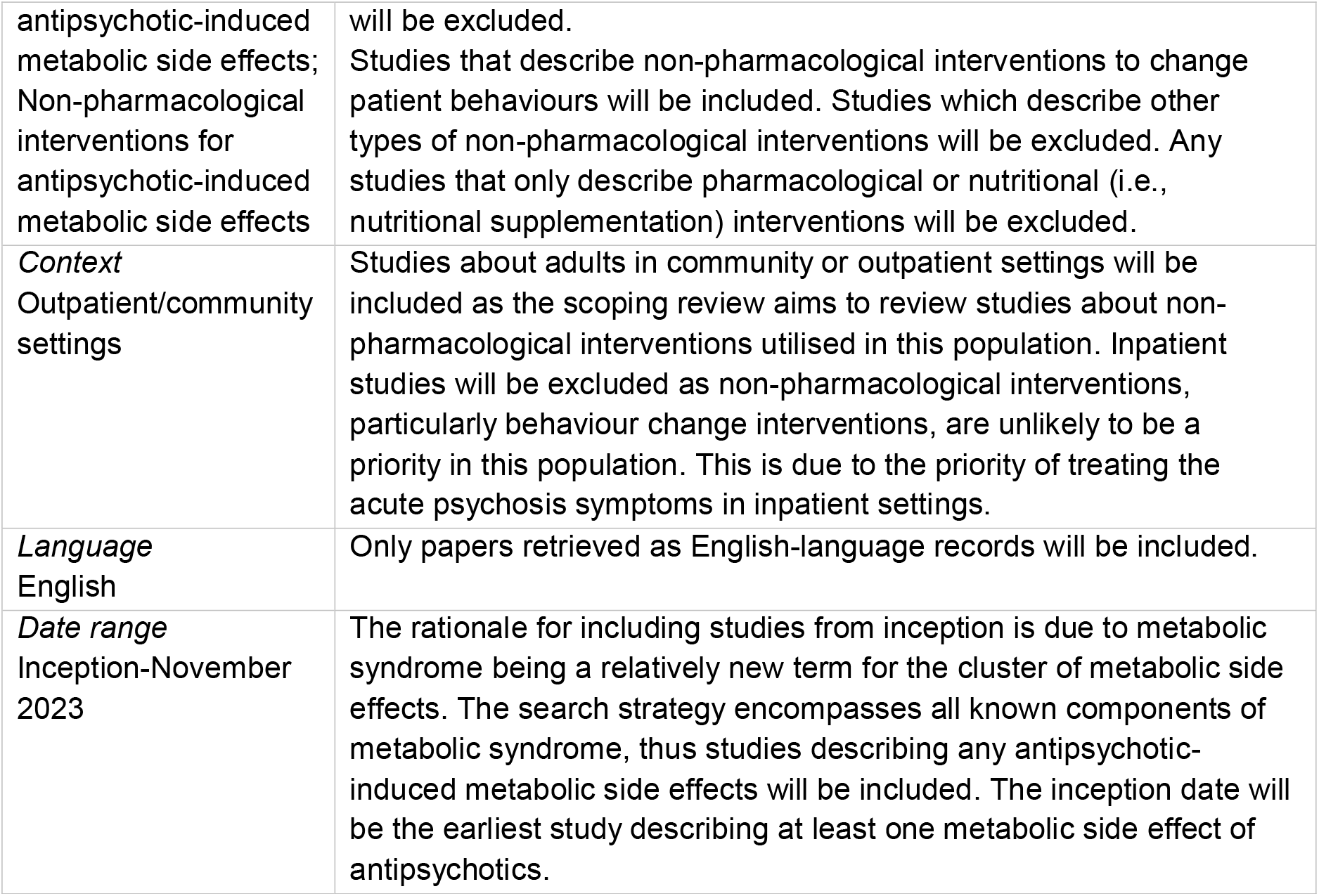
Inclusion and exclusion criteria for studies.

### 4.5 Information sources

The following bibliographic databases will be searched in November 2023 from inception to the present:

- Medline (OVID interface)
- The Cochrane Library (Cochrane Reviews)
- CINAHL (EBSCO interface)
- APA PsychInfo (EBSCO interface) (for behavioral sciences and mental health)
- Embase (OVID interface)
- Emcare (OVID interface)

The following databases will be used for supplementary database searches:

- Scopus (Elsevier)
- Epistemonikos.

One author will perform all searches. These sources will be searched using combinations of relevant search terms, adapted to the search particulars of each database. It is noted that by limiting the search to English language only, this may result in bias in the results towards English language speaking countries.

### 4.6 Screening procedure

#### 4.6.1 Title and abstract screening

All identified records will be exported into EndNote reference management software and duplicates will be removed. Titles and abstracts will be screened for inclusion using Rayyan software. Rayyan is an application designed to facilitate the screening process for systematic reviews, scoping reviews, and other literature review projects.

A screening form (Appendix 3) will be piloted on the first 50 citations to test both the criteria and reviewer agreement. After this stage, the remaining citations will be screened in Rayyan.

The primary reviewer will undertake the screening process to identify relevant studies. Where it is not clear if the eligibility criteria are met, items will automatically pass through to the full-text screening stage. Definite non-relevant sources will be excluded. To assure quality in title and abstract screening, two secondary reviewers will screen a 10% sample each. The inclusion/exclusion criteria will be adjusted as necessary. The research team will meet to discuss and resolve any discrepancies. Agreement rate and Cohen kappa coefficients will be calculated to measure inter-rater reliability.

Upon completion of the total title and abstract screening, the research team will meet to reach an agreement regarding which sources to include in the final set of eligible studies. Reasons for exclusion will be noted and reported in the manuscript.

#### 4.6.2 Full text screening

Full texts will be obtained for articles meeting the eligibility criteria. Study authors will be contacted as required for additional information. The primary reviewer will complete the full text screening using the same eligibility criteria for title and abstract screening (see Table 1) to determine final inclusion. Following the full-text screening, studies recommended for exclusion will be reviewed by a second researcher to ensure consistency in the application of exclusion criteria. Disagreements will be resolved by discussion or referral to a third reviewer. Excluded studies will be recorded along with the reason(s) for exclusion.

The final search results will be reported in a PRISMA-ScR flow diagram(39) to demonstrate the different phases of the review decision process: indicating the search results; removing duplicates; screening; selecting and retrieving studies for inclusion; clarifying reasons for exclusion during the full-text review; and the final presentation of the results.

### 4.7 Data charting

Key characteristics from all included articles will be extracted into a pre-defined Microsoft Excel data extraction form by the primary reviewer (see appendix 4). The data extraction form will be piloted with the first five studies and will be modified as necessary. A second researcher will complete a 10% check after completion of data charting to assure quality in data extraction. Study quality will not be assessed as the objective of the review is to identify gaps in the literature. The PRISMA-ScR checklist guidance pertaining to the data charting process and reporting sources of evidence will be followed(40,41).

### 4.8 Data synthesis

The charted data will be summarised and presented in a narrative synthesis to describe what is known about the similarities, differences, and relationships within the data, and to synthesise ideas and theories. This will be supported with the presentation of results in a diagram to demonstrate the distribution of studies by origin, area of intervention, and research methods. Results will be collated as they address the research question and objectives. Specifically, what is known about the following will be mapped and described:

- The target population.
- The modifiable and non-modifiable determinants associated with the development of antipsychotic-induced MSEs in the target population.
- The preferred context for delivery for a non-pharmacological intervention to address the behaviour changes associated with antipsychotic-induced MSEs.

### 4.9 Critical appraisal of individual sources of evidence

A formal quality assessment of each study will not be undertaken. Instead, the richness of the data will be categorised in terms of how well data addresses the research questions(42,43).

### 4.10 Consultation process

As per the guidance set out by JBI(28), consultation will be built into the review. At the end of the review, consultation will take place via interviews with stakeholders (comprising patients and family members/primary carers) to gain their views on the modifiable and non-modifiable determinants, and the feasibility of the preferred context for delivery of a behaviour change intervention.

### 4.11 Patient and Public Involvement

Experts by experience (n=2) and personal consultees (n=2) have been involved from the outset to help shape the focus of the research by identifying areas required to be studied to help define and prioritise the research question and objectives. These stakeholders described the status quo as “wholly inadequate” and the need to depart from a ‘watch and wait’ approach to managing antipsychotic-induced MSEs was raised as a priority. Preventative action needs to be delivered at the point of antipsychotic prescribing and not when MSEs develops, and this needs to be empirically developed in partnership with the target audience.

A Patient and Public Involvement (PPI) group (n=6) will be convened to help the research team understand the results of this scoping review. PPI will assist the research team to map the behavioural determinants this scoping review finds to the Theoretical Domains Framework(44). We will adopt the PPI model used in the CompreHensive geriatrician-led Medication Review (CHARMER)(45) which authentically embeds PPI into the research team, and we will evaluate PPI involvement using the Public Involvement Impact Assessment Framework (PiiAF)(46).

## Supporting information

Supplemental file 1 - PRISMA-ScR Checklist

Supplemental file 2 - Search strategy

Supplemental file 3 - Screening form template

Supplemental file 4 - Data extraction tool

## Data Availability

All data produced in the present study are available upon reasonable request to the authors.

## 5.0 ETHICS AND DISSEMINATION

Ethical approval is not required for this scoping review as it will involve secondary analysis of publicly available, anonymised data. The review will not involve any research participants external to the research at any point. PPI will be invited to join the research team and will not be research participants.

Findings of the scoping review will be disseminated through professional networks, conference presentations and publication in a peer-reviewed scientific journal. The PPI group will also advise on additional places share the review findings.

