## Supplemental file 2 - Search strategy for "What is known about the determinants of developing antipsychotic-induced metabolic syndrome and interventions to address them for community dwelling adults: a scoping review protocol"

**Appendix 2**

**MEDLINE search strategy**

| Ovid MEDLINE(R) ALL <1946 to November 10, 2023> | | |
| --- | --- | --- |
| 1 | exp Metabolic Syndrome/ | 38762 |
| 2 | exp *Weight Gain/de, eh, ph | 5084 |
| 3 | exp *Obesity/ci, co, di, dh, eh, et, me, mo, nu, ph, pc, px, rh, th | 82446 |
| 4 | exp *Body Weight/ae, de, eh, ph, pc, px | 41789 |
| 5 | exp *Hypercholesterolemia/ci, co, di, dh, eh, et, me, mo, nu, pc, px, rh, th | 6435 |
| 6 | exp *Cardiovascular Diseases/ci, co, di, dh, eh, et, me, mo, nu, ph, pc, px, th | 1041851 |
| 7 | exp *Cardiovascular Abnormalities/ci, co, di, dh, eh, et, me, mo, ph, pc, px, rh, th | 66009 |
| 8 | exp *Heart Diseases/ci, co, di, dh, de, eh, et, me, mo, nu, ph, pc, px, rh, th | 487963 |
| 9 | exp *Diabetes Mellitus/ci, di, dh, ed, eh, et, me, mo, nu, ph, pc, px, rh, th or Diabetes Mellitus, Type 2/ci, co, di, dh, eh, et, me, mo, nu, pc, px, rh, th | 192592 |
| 10 | exp *Dyslipidemias/ci, co, di, dh, eh, et, me, mo, nu, pc, px, rh, th | 22578 |
| 11 | exp *Lipids/ae, ph, th | 32969 |
| 12 | exp *Hyperglycemia/ci, co, di, dh, eh, et, me, mo, nu, ph, pc, px, rh, th | 11916 |
| 13 | exp *Blood Glucose/ae, de, me, ph | 30340 |
| 14 | exp *Glucose/ae, me, ph, th | 64887 |
| 15 | exp *Hypertension/ae, ci, co, di, dh, de, eh, et, me, mo, nu, ph, pc, px, rh, th | 84108 |
| 16 | exp *Body Mass Index/ | 23520 |
| 17 | 1 or 2 or 3 or 4 or 5 or 6 or 7 or 8 or 9 or 10 or 11 or 12 or 13 or 14 or 15 or 16 | 1452623 |
| 18 | metabolic syndrome.tw. | 62482 |
| 19 | diabet*.tw. | 782629 |
| 20 | lipid*.tw. | 598090 |
| 21 | dyslipid?emia*.tw. | 43925 |
| 22 | hyperlipid?emia*.tw. | 32547 |
| 23 | hypercholesterol*.tw. | 39165 |
| 24 | blood pressure.tw. | 337189 |
| 25 | hypertension.tw. | 451790 |
| 26 | blood glucose.tw. | 87700 |
| 27 | hyperglyc?emi*.tw. | 74621 |
| 28 | weight gain.tw. | 75645 |
| 29 | obes*.tw. | 381292 |
| 30 | overweight.tw. | 88880 |
| 31 | BMI.tw. | 195614 |
| 32 | body mass index.tw. | 238998 |
| 33 | cardiovascular.tw. | 552338 |
| 34 | heart.tw. | 936341 |
| 35 | 18 or 19 or 20 or 21 or 22 or 23 or 24 or 25 or 26 or 27 or 28 or 29 or 30 or 31 or 32 or 33 or 34 | 3288257 |
| 36 | 17 or 35 | 4022358 |
| 37 | exp Antipsychotic Agents/ | 129123 |
| 38 | antipsychotic*.tw. | 46603 |
| 39 | (amisulpride or aripiprazole or asenapine or clozapine or iloperidone or lurasidone or olanzapine or paliperidone or quetiapine or remoxipride or risperidone or sertindole or ziprasidone or zotepine or benperidol or chlorpromazine or droperidol or flupentixol or fluphenthixol or fluphenazine or haloperidol or levomepromazine or loxapine or mesoridazine or pericyazine or perphenazine or pimozide or sulpiride or thioridazine or thiothixine or trifluperazine or zuclopenthixol).tw. | 71449 |
| 40 | 37 or 38 or 39 | 158960 |
| 41 | exp Psychotic Disorders/ | 59081 |
| 42 | exp Schizophrenia/ | 115901 |
| 43 | exp Bipolar Disorder/ | 45498 |
| 44 | 41 or 42 or 43 | 194133 |
| 45 | (psychos* or psychotic*).tw. | 220911 |
| 46 | schizophreni*.tw. | 139703 |
| 47 | bipolar disorder*.tw. | 34550 |
| 48 | 45 or 46 or 47 | 352675 |
| 49 | 44 or 48 | 403020 |
| 50 | 36 and 40 and 49 | 7150 |
| 51 | limit 50 to "all adult (19 plus years)" | 3730 |
| 52 | limit 51 to english language | 3488 |
| 53 | (address or autobiography or bibliography or biography or case reports or clinical conference or clinical trial, veterinary or clinical trial protocol or comment or congress or consensus development conference or consensus development conference, nih or dataset or dictionary or directory or editorial or english abstract or electronic supplementary materials or "expression of concern" or festschrift or government publication or guideline or interactive tutorial or interview or introductory journal article or lecture or legal case or legislation or letter or news or newspaper article or observational study, veterinary or overall or patient education handout or periodical index or personal narrative or portrait or practice guideline or randomized controlled trial, veterinary or technical report or video-audio media or webcast).pt. | 6288578 |
| 54 | 52 not 53 | 2664 |
