## Supplemental file 3 - Screening form template for "What is known about the determinants of developing antipsychotic-induced metabolic syndrome and interventions to address them for community dwelling adults: a scoping review protocol"

**Appendix 3**

**Screening form template**

| **No.** | **Link to full text** | **Title** | **Author** | **Journal** | **Year** | **MSE Topic** | **Non-modifiable determinants** | **Modifiable determinants** | **Preferred implementation of intervention** | **Inpatient/**  **Outpatient** | **Relevant Y/N** | **Evaluation** |
| --- | --- | --- | --- | --- | --- | --- | --- | --- | --- | --- | --- | --- |
| 1 |  |  |  |  |  |  |  |  |  |  |  |  |
| 2 |  |  |  |  |  |  |  |  |  |  |  |  |
| 3 |  |  |  |  |  |  |  |  |  |  |  |  |
| 4 |  |  |  |  |  |  |  |  |  |  |  |  |
| 5 |  |  |  |  |  |  |  |  |  |  |  |  |
| 6 |  |  |  |  |  |  |  |  |  |  |  |  |
| 7 |  |  |  |  |  |  |  |  |  |  |  |  |
| 8 |  |  |  |  |  |  |  |  |  |  |  |  |
| 9 |  |  |  |  |  |  |  |  |  |  |  |  |
| 10 |  |  |  |  |  |  |  |  |  |  |  |  |
| 41 |  |  |  |  |  |  |  |  |  |  |  |  |
| 42 |  |  |  |  |  |  |  |  |  |  |  |  |
| 43 |  |  |  |  |  |  |  |  |  |  |  |  |
| 44 |  |  |  |  |  |  |  |  |  |  |  |  |
| 45 |  |  |  |  |  |  |  |  |  |  |  |  |
| 46 |  |  |  |  |  |  |  |  |  |  |  |  |
| 47 |  |  |  |  |  |  |  |  |  |  |  |  |
| 48 |  |  |  |  |  |  |  |  |  |  |  |  |
| 49 |  |  |  |  |  |  |  |  |  |  |  |  |
| 50 |  |  |  |  |  |  |  |  |  |  |  |  |
